# Low-cost rare variant detection for population scale genetic screening

**DOI:** 10.64898/2026.07.07.26357001

**Authors:** Mads Cort Nielsen, Caroline M. Junker Mentzel, Ulrik Kristoffer Stoltze, Christian Munch Hagen, Marie Bækvad-Hansen, Anna Byrjalsen, Lone Sunde, Alberte Aspaas Lundquist, Allan Meldgaard Lund, Jacob Tfelt-Hansen, Tania Masmas, Erik Sørensen, Ole Birger Vesterager Pedersen, Christian Erikstrup, Sisse Rye Ostrowski, DBDS Genomic Consortium, Henrik Hjalgrim, Mette Nyegaard, Kjeld Schmiegelow, Thomas van Overeem Hansen, Karin Wadt, Jonas Bybjerg-Grauholm, Simon Rasmussen

## Abstract

Genetic screening for rare pathogenic variants facilitates early detection and prevention of disease manifestations in medically actionable disorders, but sequencing costs limit widespread use. We introduce DoBSeq, a low-cost, high-throughput screening framework for detecting rare, single-nucleotide variants and indels. The framework includes: extraction of DNA from dried blood spots used in neonatal screening, automation of two-dimensional DNA pooling and library preparation, high-depth targeted sequencing using a 582-gene custom panel, and a probabilistic model to assign rare pathogenic variants to individuals. Benchmarked against whole-genome sequencing across 582 genes in a batch of 576 individuals, the framework detected 95% of all variants and recovered all clinically relevant pathogenic single-nucleotide variants in American College of Medical Genetics and Genomics (ACMG) actionable genes. Applied to 2304 anonymised blood donors, it yielded variant frequencies consistent with existing population estimates. At a sample cost of 29 USD, including 11 USD running costs, this framework provides a cost-efficient approach to population-level genetic screening.

## INTRODUCTION

Rare diseases (RDs) affect approximately 6% of the European population^1^ and are associated with high mortality, morbidity, disability, and healthcare costs^2,3^. While population screening could help address these issues, traditional biochemical assays are unsuitable for the majority of RDs due to their low prevalence or lack of sufficiently sensitive and specific biomarkers^4^. Since 70% of RDs have a genetic cause^5,6^, DNA sequencing offers a promising approach to test for these conditions in a simultaneous way^7^.

Many large-scale studies incorporating DNA sequencing are underway, most of which are based on next-generation sequencing of whole exomes or whole genomes^8–10^. These studies provide insight into the prevalence and penetrance of rare disease-associated variants in the broader population. However, the cost of applying individual-level sequencing across entire populations remains prohibitive. To address this, the Danish PREDiSPOSED project, a population-based study of diagnostic sequencing, employs double-batched sequencing (DoBSeq), an overlapping pooled-sequencing strategy in which individual DNA samples are arranged in a matrix and pooled twice, once along each axis. In this design, any row pool and column pool share one individual. Variants detected in exactly one row pool and one column pool can therefore be uniquely assigned to the individual at their intersection^11^. Matrix-unique variants are referred to as private variants throughout the study.

This design reduces costs by minimizing library-kit usage, sample handling after DNA pooling. In our previous work^11,12^, we evaluated the method in pilot batches of 100 individuals arranged in 10×10 matrices and achieved rare-variant detection sensitivities comparable to individual sequencing for clinically relevant variants. Because the number of sequencing libraries scales with the number of pools rather than the number of individuals, larger matrices substantially reduce per-sample library-preparation cost, motivating scaling beyond the 10×10 configuration. However, increasing the number of samples per pool increases the complexity of sample handling and variant calling, requiring specialized laboratory and bioinformatic workflows, as well as additional validation.

One issue of scaling to larger batch sizes is that assignment of variants in the DoBSeq matrices becomes limited by technical variation introduced during DNA pooling and library preparation. Individual DNA contributions are never perfectly equimolar, even after thorough quantification and normalization. This variation can be amplified further during library preparation, resulting in significant variation in the distribution of allele-supporting reads and the estimated allele fractions. As a result, variants can be missed in one of the two pools containing DNA from the carrier. This incomplete detection prevents correct assignment and leads to false-negative results that become more frequent with increasing matrix size and resulting DNA inequality. The two-dimensional pooling design, however, provides two independent observations per individual. A variant missed in one pool can still be identified in reads in the other dimension. Probabilistic approaches that integrate read-level evidence and allow pool-specific variation in the expected fraction of variant supporting reads have been developed for variant calling in pooled sequencing^13,14^. Extending this approach to the two-dimensional DoBSeq design offers a possible way to use this redundancy and recover variants otherwise missed.

Here, we present a refined and extended framework for performing DoBSeq using a 582-gene panel in batch sizes of 576 individuals arranged in a 24×24 matrix. The framework includes automated DNA extraction and pooling of DNA from dried blood spots used in neonatal screening, along with an optimized library-preparation protocol that supports the increased sequencing depth required at this scale. To improve detection sensitivity and individual-level assignment, we developed a Bayesian probabilistic model that leverages the repeated sequencing of each individual across a row pool and a column pool. The model combines per-pool read evidence with initial variant calls to estimate the probability that a candidate individual is the unique carrier of a given variant. We validated the framework in a childhood cancer cohort with matched whole-genome sequencing (WGS) data, achieving a sensitivity of 95% for all evaluated variants and assigning the majority of pathogenic variants in genes included in the American College of Medical Genetics and Genomics Secondary Findings v3.2 list (ACMG SF v3.2). We then applied the framework to four 24×24 matrices comprising a retrospective cohort of 2,304 anonymised blood donors, recovering known population-genetic allele frequencies at a fraction of the per-sample cost of individual sequencing.

**Figure 1.**
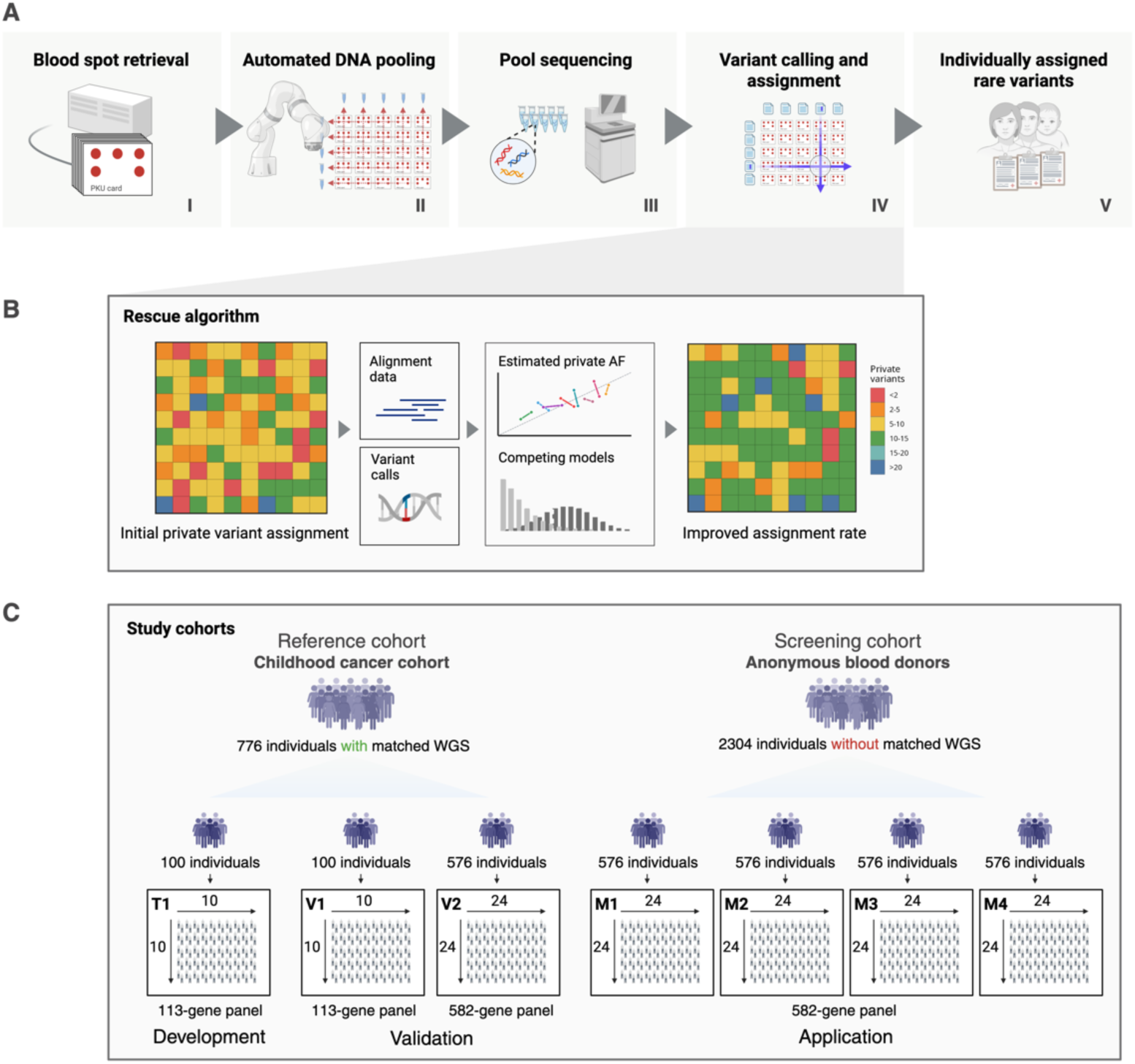
Overview of the DoBSeq framework, rescue algorithm and study cohorts. **(A)** (I) individual dried blood spot samples are retrieved and DNA is extracted from 3.2-mm disc excised blood-spots. (II) Using an automated workflow, DNA is normalized to equimolar concentrations, arranged in a matrix format, and pooled along each dimension so that each individual’s DNA is present in exactly two pools. (III) Next, library preparation and deep target capture sequencing are performed on the pooled DNA samples. (IV) A specialized computational pipeline processes the sequencing data through alignment, variant calling, filtering, and assignment of variants to individuals. (V) This framework enables linking rare variants to their individual carriers. (**B**) A rescue algorithm recovers private variants by integrating initial variant calls and raw alignment data in a Bayesian probabilistic model. (**C**) The framework was developed and validated using a reference cohort of 776 individuals from a childhood cancer cohort with matched WGS data. The reference cohort comprised a 10×10 training matrix (T1) and two held-out evaluation matrices (V1, 10×10; V2, 24×24). T1 and V1 were sequenced using a commercial 113-gene panel, whereas V2 was sequenced using a 582-gene custom panel. The validated framework was then applied to a screening cohort of 2,304 anonymized adult blood donors without matched WGS data, arranged in four 24×24 matrices (M1–M4) and sequenced using the same 582-gene custom panel.

## RESULTS

### Study design and cohorts

In the process of developing, validating, and applying the extended DoBSeq framework, we included two separate cohorts that were distributed across seven pooling matrices (**Figure 1**). The method development included a previously published childhood cancer reference cohort, arranged in a 10×10 training matrix (T1) and a 10×10 validation matrix (V1), both sequenced with a 113-gene hereditary cancer panel. To validate the framework at the larger batch size, an additional 576 individuals from the same reference cohort were arranged in a 24×24 matrix (V2) and sequenced using a custom 582-gene panel that combined the 113-gene hereditary cancer panel with all genes from the ACMG SF v3.2 list, along with additional rare disorder-associated genes (see **Table S1**). The framework was then applied to a screening cohort of 2,304 anonymous adult blood donors, arranged in four independent 24×24 matrices (M1–M4) and sequenced with the same 582-gene panel.

### Development of a variant recovery model

To improve the sensitivity of variant assignment and utilise the duplicate sequencing information from each individual, we developed a Bayesian probabilistic model to evaluate variants called in only a single dimension (see **Figure 2A-C**). The model extracts total coverage and variant supporting read-depth at every called position across all pools and compares two competing models at each site. A reference model (M0) that assumes that the variant-supporting reads arose from noise, due to sequencing and alignment errors, and a carrier model (M_1_) that assumes that the variant-supporting reads originate from one specific individual. The evidence for each candidate is quantified using a Bayes factor, and the probability of an individual being the sole carrier is computed as the joint probability of that individual carrying the variant and all others not carrying it. Pipetting and DNA measurement uncertainty can cause the estimated fraction of variant-supporting reads to vary among individuals. The model accounts for this by estimating individual-specific frequencies from the initial assigned variants using an empirical Bayesian approach. (See **Figure S1** for an overview of the algorithm).

### Probabilistic rescue model improves rare variant assignment sensitivity in pool sequencing data

The rescue model was developed and adjusted on the 10×10 training matrix (T1) (See **Figure 1C**). As a baseline, we evaluated a Z-score approach that quantifies the distance between candidate pool coverage and the distribution of non-candidate pools. Both approaches were then tested on a completely separate held-out 10×10 matrix (V1) (see **Figure S2**) and the 24×24 validation matrix (V2). The Bayesian model and Z-score approach had a similar high performance in terms of ROC-AUC (receiver operating characteristics area under the curve), with the Bayesian model performing slightly better (0.976 vs. 0.97) in classifying variants as assignable or not (see **Figure 2D**). Using a default (0.5) threshold for the Bayesian model and a F1-optimised threshold for the Z-score approach, the models both achieve a sensitivity increase (Bayesian: +2.1%, Z-score:+3.6%), by recovering 234 (228/6, SNV/indels) and 262 (250/12, SNV/indels) variants respectively, compared to only using variants called in both dimensions (see **Figure 2E-F**). While the Bayesian method increased precision (+2.2%) and F1-score (+2.1%), the Z-score substantially reduced both metrics (Precision: -33.7%, F1-score: -21%), indicating that the model was less robust to the change in matrix size. Based on the more stable performance improvements, the Bayesian method was implemented as a default addition to the DoBSeq framework.

**Figure 2.**
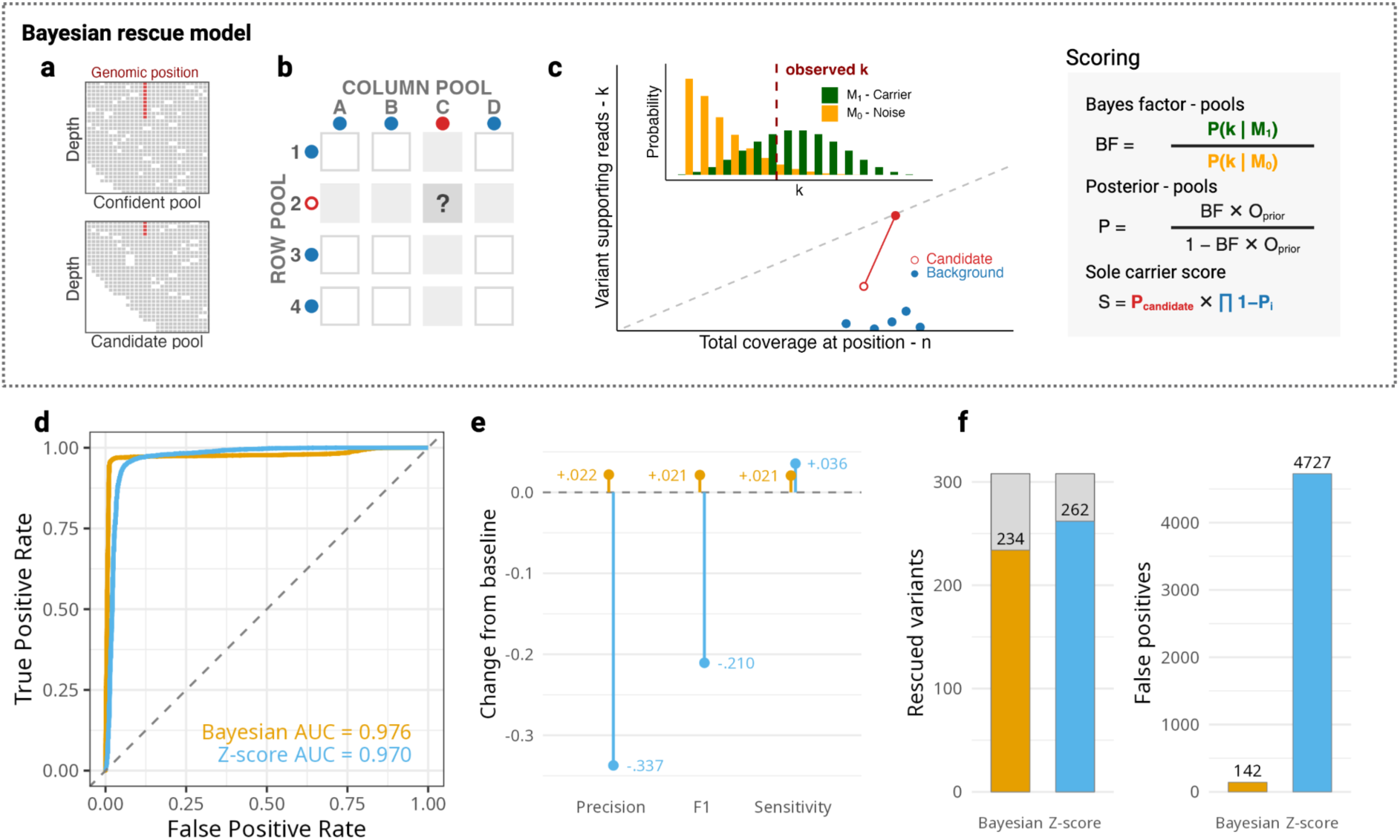
Overview of the variant rescue algorithm and performance. (**A**) The algorithm takes confident variant calls and alignment data with alternative allele support (red) from the primary workflow. (**B**) At each genomic position where a variant has been called in a single pool only (filled red circle), each pool in the opposite dimension is evaluated as a candidate carrier (open red circle). Each pool intersection corresponds to a distinct individual, with the evaluated candidate marked “?”. (**C**) For each candidate pool, two competing empirical Bayes models are compared: a reference noise model (M0, yellow) and an individual-specific carrier model (M1, green). The scatter plot shows variant-supporting reads vs. total coverage across all pools at the specific genomic position (candidate pool in red, background pools in blue). The histogram insert shows the probability of observing k variant-supporting reads under each model, given the total coverage n. The observed k (dashed line) is evaluated with both models to compute the Bayes factor. A Bayes factor per pool is converted to a posterior probability, and a final sole-carrier score for the specific individual is computed as the joint probability that the candidate pool carries the variant and all others do not. (**D**) Model performance shown as ROC-AUC using all variants in the 24×24 validation matrix for the Bayesian variant rescue model (orange) and the Z-score baseline model (blue). (**E**) Model effect on the performance in assigning private variants to individuals in the validation matrix. Changes in performance are shown as rate changes relative to the primary workflow output without a rescue step. (**F**) Number of recovered variants out of the total recoverable variants and the total number of false positive variant assignments for the Bayesian variant rescue model (orange) and the Z-score baseline model (blue). Grey bars indicate the remainder of the recoverable set.

### Validation of the extended DoBSeq framework using a cohort of 576 individuals

To evaluate whether the extended framework reliably detects and assigns rare variants at the 24×24 scale, we validated it against matched WGS data from the 24×24 reference matrix (V2). To mimic a neonatal screening setting, defrosted whole-blood was spotted onto filter paper and DNA was extracted. Individual DNA samples were arranged in a 24×24 matrix, and the pooled samples were processed through the DoBSeq framework using a custom panel of 582 genes (see **Figure 3**). The automated pooling and optimized library preparation produced libraries with increased complexity compared to the 10×10 pilot, with balanced per-individual contributions and uniform coverage across the 48 pools (see **Figure S3**).

Across the target regions, WGS identified 176,302 distinct variants, of which 167,985 (95%) were detected by the DoBSeq framework (see **Figure 3**). Allele frequencies estimated from the pooled data were concordant with the WGS reference for multi-carrier variants (Spearman ρ = 0.98, MAE = 0.002; **Figure S4**). The primary application of DoBSeq, however, is detecting private variants, which can be unambiguously assigned to that individual. To evaluate assignment performance, we identified all private variants from the individual WGS call sets and compared these to the DoBSeq-assigned variants. Considering all private variants, regardless of their predicted biological consequence, DoBSeq correctly assigned 6,450 of 7,087, achieving a sensitivity of 91% and a precision of 86.5%.

We next assessed performance on clinically relevant variants, defined as predicted high-confidence loss-of-function variants and ClinVar-annotated likely-pathogenic or pathogenic variants. Of these, 259 were private, and the framework correctly assigned 225 (168/184 SNVs, 57/75 indels; sensitivity 86.9%, precision 69.0%). Additionally, a panel of clinical experts reviewed these variants along with all variants of unknown significance in 81 genes included in the ACMG SF v3.2 guideline^15^. Of the 23 variants reviewed, 19 were correctly detected and assigned.

For a subset of samples (n=104), DNA concentrations from the dried blood spot cards were too low to allow equimolar contributions to the pools, and these individuals were therefore expected to be under-represented in the sequencing data. After excluding these samples, performance improved across all metrics: 5,450 of 5,827 private variants were correctly assigned (sensitivity 93.5%, precision 87.1%); 190 of 216 private clinically relevant variants were assigned (140/148 SNVs, 50/68 indels; sensitivity 88%, precision 74.5%); and 16 of 18 expert-reviewed variants were correctly detected and assigned (**Figure S5**). The remaining two false-negative reviewed variants, a *BRCA2* (MIM: 600185) and a *WT1* (MIM: 607102) indel, were not called in any of the pools, despite their genomic positions being covered and their carriers having multiple other variants assigned (**Figure S6**). The *WT1* indel overlapped with another variant assigned to the same individual, but the *BRCA2* had zero alternative coverage. The largest sensitivity gains from the rescue model were in samples with the lowest pre-pooling DNA concentrations (Spearman’s ρ = -0.39 between concentration and sensitivity gain, one-sided p < 2.2 × 10⁻¹⁶) (**Figure S7**).

**Figure 3.**
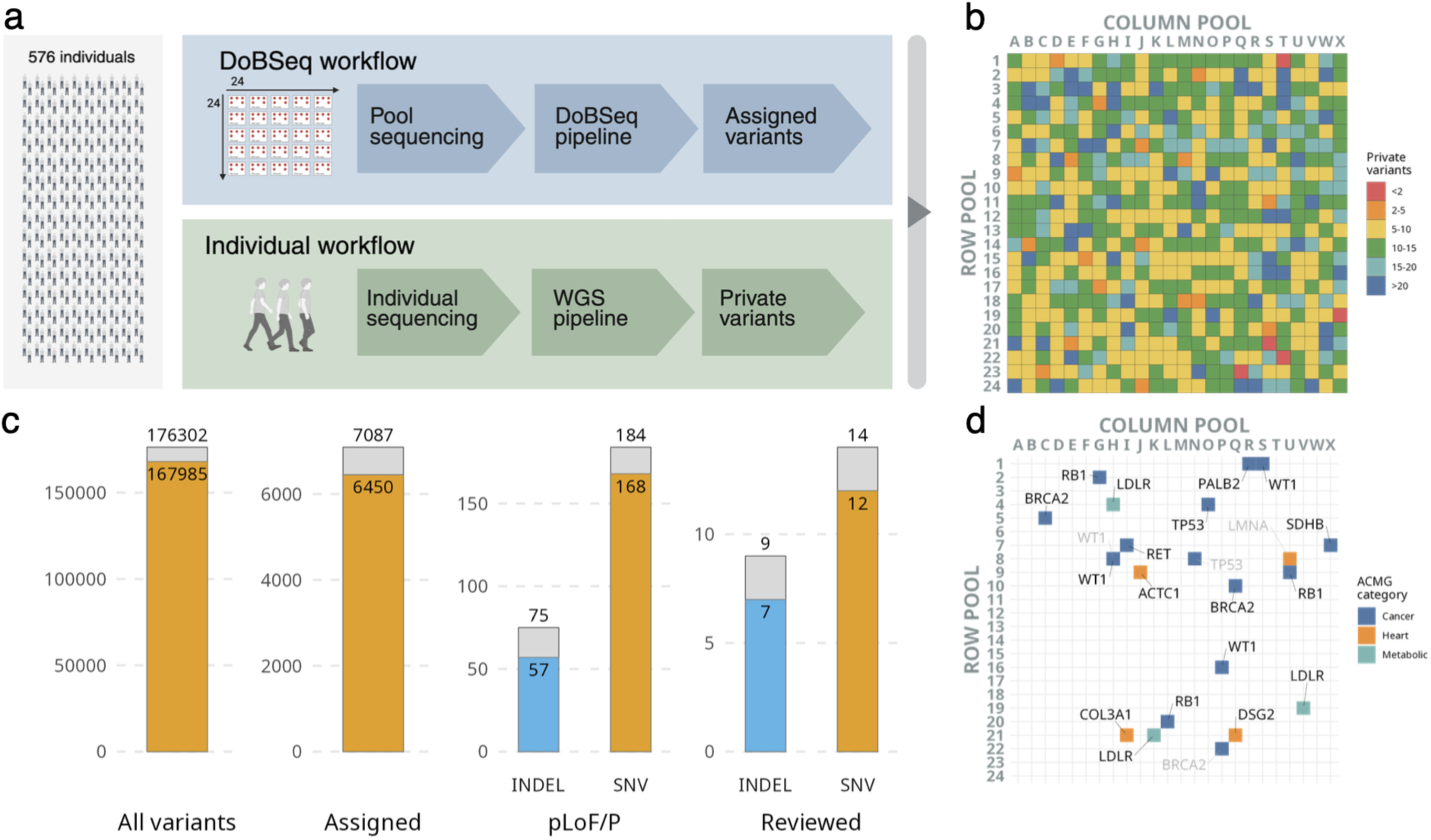
Validation of the DoBSeq framework on a batch size of 576 individuals. (**A**) The framework was validated using a cohort of 576 childhood cancer patients via two parallel approaches: DoBSeq screening with a 24×24 matrix format targeting 582 rare disease-associated genes (I), and individual germline whole genome sequencing (WGS) with data subset to the DoBSeq panel regions and filtered for private variants (II). (**B**) Heatmap of the 24×24 matrix showing the number of private variants per individual (WGS), with color indicating count intervals. (**C**) Bar charts comparing DoBSeq output (orange) against the individual workflow (grey) at successive filtering stages: all distinct variants (private and common) across the dataset, assigned variants versus private variants, the subset annotated as predicted loss-of-function or ClinVar pathogenic/likely pathogenic (pLoF/P), and expert-reviewed variants within ACMG genes. (**D**) Reviewed variants positioned at their corresponding individual’s location in the 24×24 matrix and colored by ACMG category. Variants shown in light grey were identified by individual WGS but not detected by DoBSeq.

### Variant sharing across pools showed most pLoF variants were assignable in a 24×24 matrix

The framework only assigns private variants to individuals. Consequently, when two or more individuals carried the same variant, the variant was called in multiple pools per dimension, and direct assignment was not possible. We characterized this constraint using WGS data from the reference individuals arranged in the 24×24 experimental matrix (**Figure 4A**). As expected, an increase in the number of pools with variant calls was associated with an increase in the mean number of carriers in the matrix (**Figure 4B**). As more pools were called, the number of possible row–column intersections grew combinatorially, and with it the variance in carrier counts (mean carriers at 4 positive pools: 2.01, SD 0.0863; mean carriers at 48 positive pools: 402, SD 145). The distribution of distinct variants across pool-call counts was bimodal (**Figure 4C**). The majority (57%) were singletons found in exactly two pools (one row and one column), and a second peak of 1747 variants (14%) was found in all 48 pools, spanning the full theoretical carrier range from 24 (when carriers occupy distinct rows and columns) to 576 (full matrix). The large majority (134, 84%) of predicted loss-of-function (pLoF) variants, were singletons, with only a few exceptions. As an example, the *PAH* (MIM: 612349) splice variant c.1315+1G>A, p.(?) was called in 4 pools, corresponding to 4 candidate intersections, and was carried by 2 individuals based on the reference data. Additionally, the *CFTR* (MIM: 602421) in-frame deletion p.(Phe508del) was called in 27 pools and carried by 17 individuals in the matrix. These results show that the framework’s restriction to private variants captures the great majority of pLoF variants with few but important exceptions.

**Figure 4.**
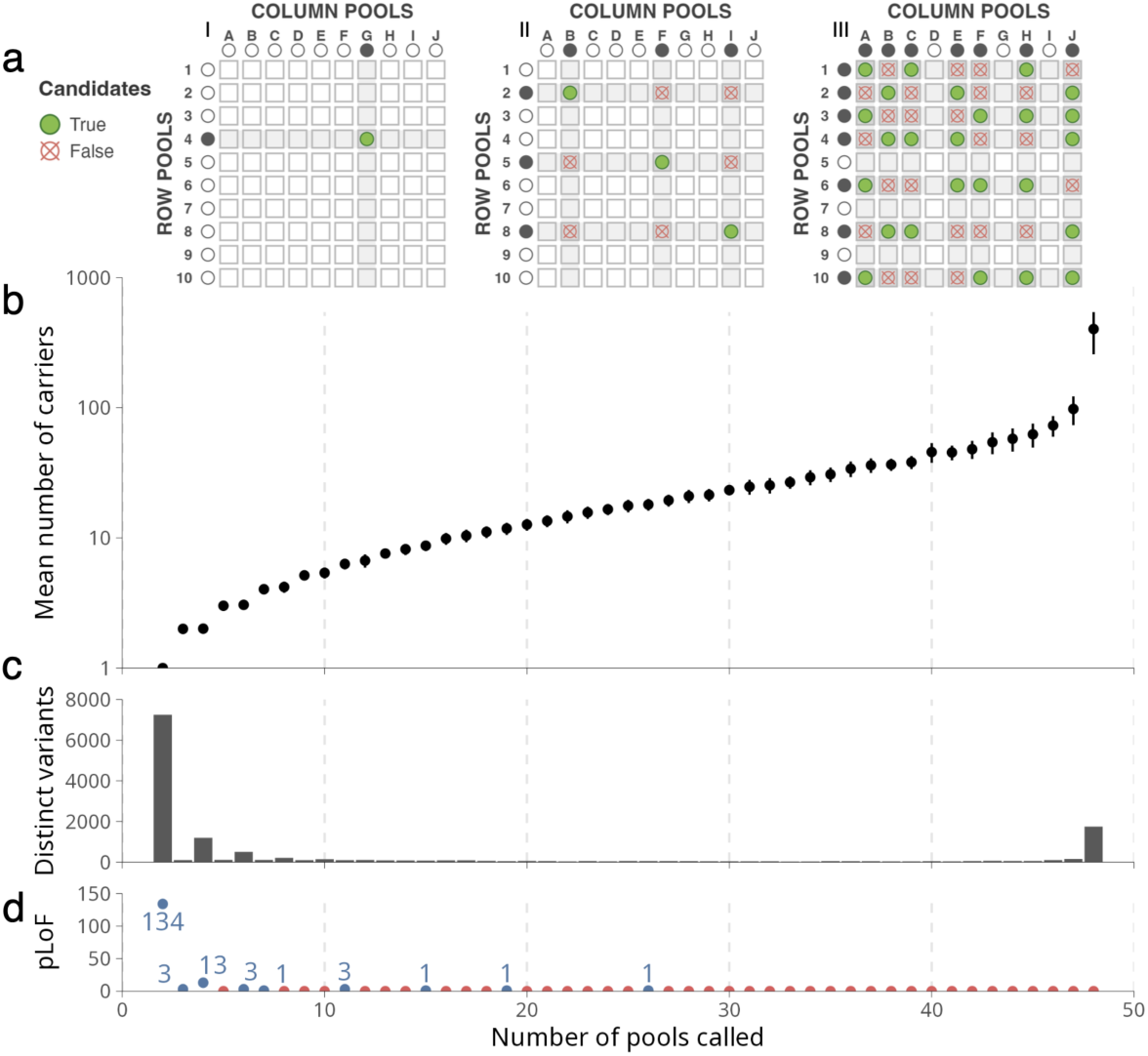
Variant sharing across pools in the 24×24 validation matrix. (**A**) Schematic illustration (shown at 10×10 for clarity; data in b–d are from the 24×24 matrix) of assignment uncertainty when a variant is called in increasing numbers of pools. Panels I, II, and III show variants called in 2, 6, and 14 pools, respectively. Black dots outside the matrices indicate positive row and column pools; grey squares mark the individuals contained in those pools. Green dots denote true candidates, and red crossed-dots denote candidate row–column intersections that do not contain a carrier (false candidates). (**B**) Mean number of true carriers per variant (determined from individual WGS data) as a function of the number of pools in which the variant was called (log₁₀ scale). Error bars show 95% confidence intervals of the mean. (**C**) Number of distinct variants at each pool-call count. (**D**) Number of predicted loss-of-function (pLoF) variants at each pool-call count. Blue dots indicate one or more pLoF variants, with the count labeled; red dots indicate zero pLoF variants.

### DoBSeq detected a *TP53* mutation mosaicism missed by individual WGS sequencing

In-depth evaluation of the validation matrix variant classification results led to the re-evaluation of the reference WGS data. Participant #O4 in the 24×24 validation matrix was included in the iCOPE project^16^ due to low-hypodiploid acute lymphoblastic leukemia (ALL) (34 chromosomes; DNA index 0.73) diagnosed in adolescence, an ALL subtype characteristically associated with Li-Fraumeni syndrome (LFS; MIM: 151623). The project undertook germline WGS with screening for rare variants in 410 cancer-related genes, yielding a normal result without reportable variants. However, upon review of false positive variant calls in the present study, the locus plot demonstrated a suspiciously clear signal for a pinpointable pathogenic missense variant in *TP53* (MIM: 191170, NM_000546.6:c.824G>A p.(Cys275Tyr), see **Figure 5B**), known to cause LFS in patients constitutionally heterozygous for the variant. In comparison to the VAFs of the patient’s other unique variants, the variant call indicated mosaicism.

Reanalysis of WGS data without VAF-filtering detected the variant in 8% [3/36] of reads. NGS panel sequencing of DNA from a blood sample collected during remission detected the variant at a VAF of 18%. In subsequent samples from the now adult patient, the variant was detected in blood in 7.7% [130/1688] of reads and in a nasal polyp at 7.8% [32/409]. However, in cultured fibroblasts and in the patient’s at-birth filter paper blood sample, the variant was not detected at 1,513 and 1,925 read-coverage, respectively. The patient remains cancer free at >15 years of follow-up, and the patient’s offspring have tested negative for the variant. These data support that the variant is somatic (or clonal) and likely isolated to the bone marrow, and the case illustrates that the relatively high-depth sequencing with DoBSeq may identify genetic variation even at low frequency, including mosaicism, that would otherwise be missed at the 30x coverage typical for WGS-based routine individual sequencing.

**Figure 5.**
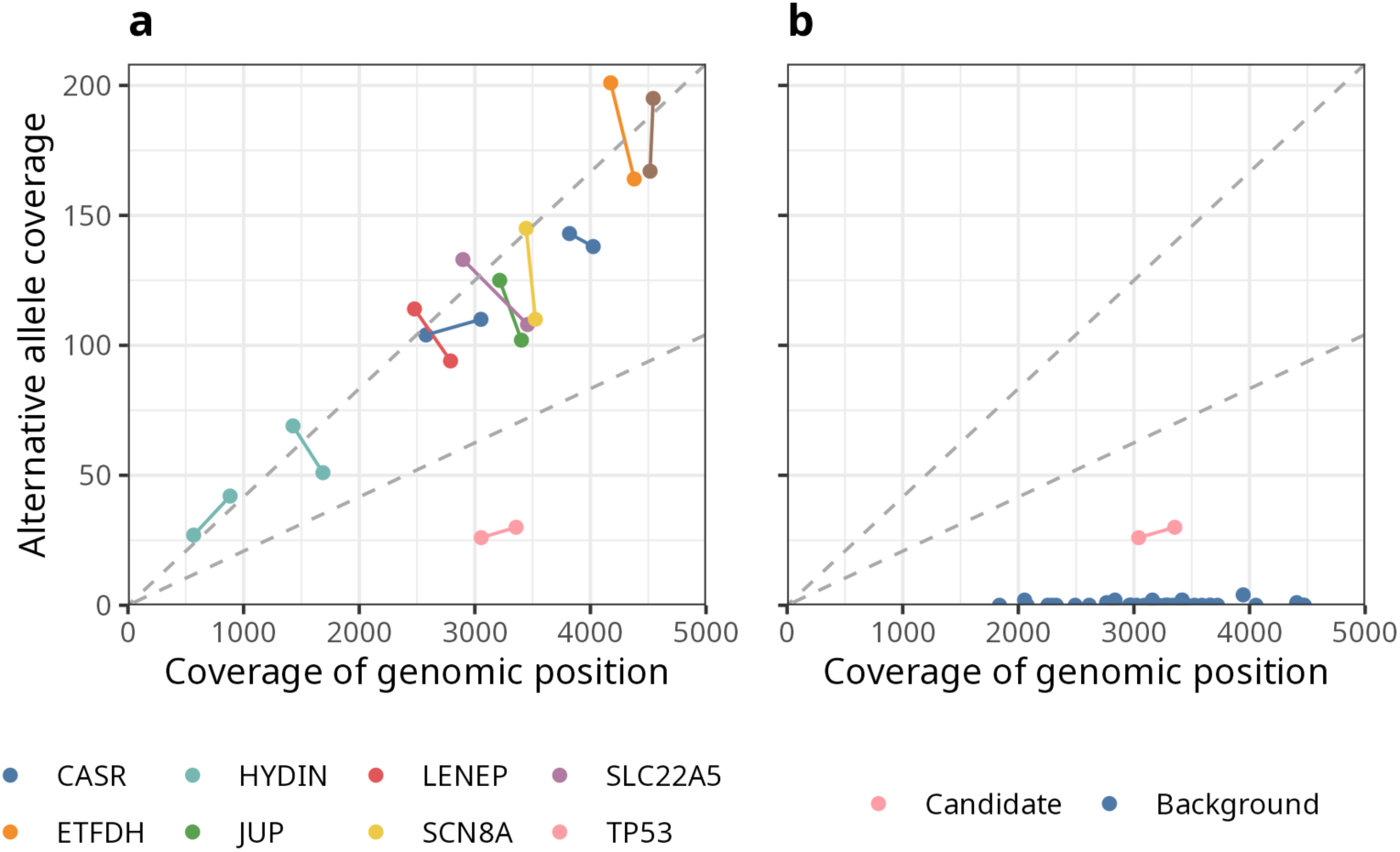
*TP53* mosaicism detected by DoBSeq. (**A**) Coverage and alternative allele coverage of all variants assigned to individual #O4 in the validation matrix. Each pair of connected dots represents one variant observed in the two pools (row and column) that uniquely identify the individual. Dashed lines indicate expected alt-allele coverage for homozygous and heterozygous variants, respectively. (**B**) Coverage and alternative allele coverage at the *TP53* variant position for the row–column pair assigned to individual #O4 (rose) compared with the same position in the remaining 46 pools (blue).

### Application to 2304 anonymous blood donors yields screening results and allele frequencies consistent with population estimates

To demonstrate the DoBSeq framework at a screening relevant scale, we applied it to the screening cohort of 2304 anonymous Danish blood donors (M1-M4). All four matrices were processed through the framework and yielded consistent results. The median number of distinct variants per pool ranged from 3897 to 4052 across matrices, and a total number of assigned variants per matrix ranged from 5910 to 6547 (**Figure 6A**). Per matrix counts of synonymous, missense, and pLoF variants were similarly comparable (synonymous: 1595–1755; missense: 2835–3074; pLoF: 137–215). Individuals had a median of 10 private variants within all target regions with comparable distributions across matrices (see **Figure S8**).

To assess the framework’s ability to capture population-level allele-frequency patterns, we compared the estimated frequencies of all detected variants in the 2304-individual cohort with those from the Danish population variant database DanMAC5^17^. This analysis was restricted to SNV coding variants with multiple carriers. We excluded singletons, for which allele frequency is fixed by the matrix design, as well as variants present in all 48 pools, for which the estimation model was uninformative. After these exclusions, 7751 variants overlapped between the two datasets. Cohort and reference allelic frequencies were concordant (Spearman ρ = 0.87, MAE = 0.0014; **Figure 6C**). A comparable trend was observed against the gnomAD non-Finnish European dataset (Spearman ρ = 0.83, MAE = 0.0019; **Figure S9**).

We next evaluated the framework’s output of clinically relevant variants. From all the detected variants in the cohort, we extracted high-confidence pLoF variants and ClinVar-annotated likely pathogenic or pathogenic (pLoF/P) variants. Of the total 1115 distinct pLoF/P variants, 848 were directly assignable (76%, 95% CI: 73-78%). Restricting the regions to only genes covered by the ACMG SF v3.2 guideline reduced the number of pLoF/P variants to 183 (131 private, 71%, 95% CI: 65-78%) (**Figure 6C**). Finally, the entire set of 183 pLoF/P ACMG SF variants was manually reviewed and classified by a panel of clinical geneticists in accordance with ACMG/AMP guidelines^18^. This resulted in a set of 37 distinct variants assigned to 42 individuals across the 4 matrices (see **Table S2**), translating to 1.8% (=42/2304, 95% CI 1.35–2.45%) being carriers of at least one pathogenic variant in an ACMG SF gene (**Figure 6E**), within the range of secondary findings rates reported by large adult genomic screening programs^19–21^.

Focusing on carrier frequencies within select conditions yielded similar results. For *BRCA1/2*, associated with hereditary breast and ovarian cancer (HBOC, MIM: 604370), the review identified 10 candidate carriers across the cohort (0.43%, 95% CI 0.24–0.8%), in line with an estimated Danish population frequency of approximately 1 in 500^22^. For Lynch syndrome (LS, MIM: 120435) genes (*MLH1* [MIM: 120436]*, MSH2* [MIM: 609309]*, MSH6* [MIM: 600678]*, PMS2* [MIM: 600259]), 3 candidate carriers were identified (0.13%, 95% CI 0.04–0.38%), within the range of the reported estimate of ∼1 in 279^23^. For familial hypercholesterolemia (FH, MIM: 143890) genes (*LDLR* [MIM: 606945]*, APOB* [MIM: 107730]*, PCSK9* [MIM: 607786]), 1 candidate carrier was identified (0.04%, 95% CI 0.002–0.24%). This is lower than Danish clinical prevalence estimates for FH^24^. However, this estimate also includes individuals without an identifiable causal variant in the three assessed genes. Four variants (*LDLR*: W23X, W66G, W556S; *APOB*: R3500Q) account for approximately 40% of FH-associated pathogenic mutations in Denmark^25^. Of these, only *APOB* R3500Q was detected in the screening cohort, with none of the three *LDLR* variants called across the four 24×24 matrices (M1-M4). However, in the 24×24 validation matrix (V2), the *LDLR* W556S variant was correctly assigned to its carrier, along with two other FH-associated *LDLR* pathogenic variants, each correctly assigned to their carriers, thereby accounting for all FH-associated pathogenic variants present in that cohort. This indicates that the framework can detect *LDLR* variants when present, and the absence in the screening cohort may be due to sampling variation.

The DoBSeq framework was used to process a total of 10 24×24 batches in this study. The resulting total per-sample cost was estimated at USD 28.5 including labor and operational work, with reagent-only cost at USD 11 per sample.

Together, these results demonstrate that the DoBSeq framework recovers cohort-level allele frequency patterns and secondary findings rates consistent with published estimates from individual sequencing at a cost-efficient per-sample cost.

**Figure 6.**
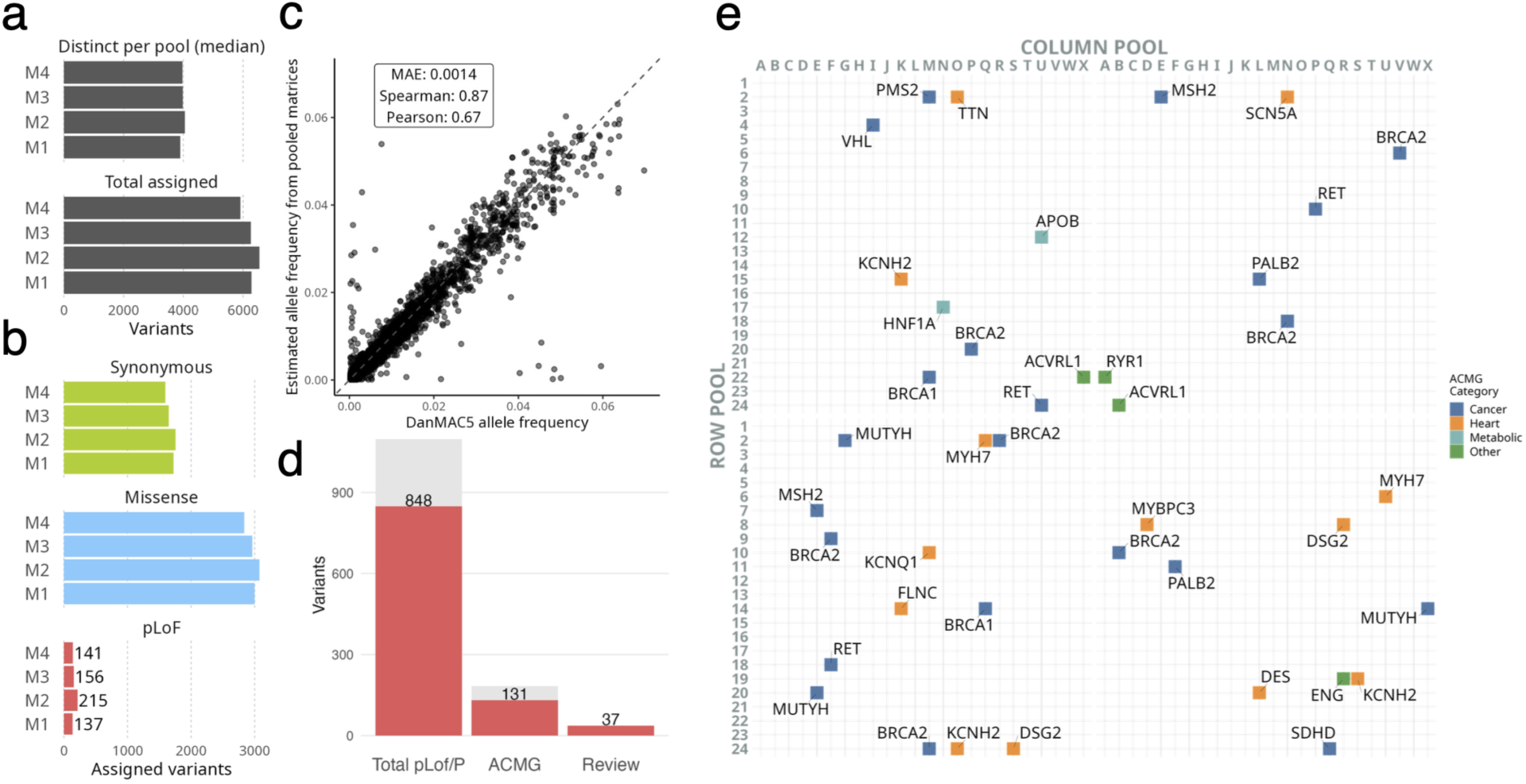
Application of the DoBSeq framework to a screening cohort of 2,304 anonymous blood donors sequenced across four 24×24 matrices. **(A)** Median number of distinct variants per pool (across the 48 pools per matrix, M1-M4) and total number of variants assigned to individuals, shown per matrix. **(B)** Per-matrix counts of assigned variants by annotation category: synonymous, missense, and predicted loss-of-function (pLoF). **(C)** Concordance between cohort-level allele frequencies estimated from the four pooled matrices and reference allele frequencies from the Danish population variant database DanMAC5. Restricted to 7,751 multi-carrier SNV coding variants present in both datasets. Statistics: mean absolute error (MAE), Spearman ρ, and Pearson r. The dashed line indicates y = x. **(D)** Clinically relevant variants in the cohort. From left to right: total distinct high-confidence pLoF and ClinVar likely-pathogenic/pathogenic variants (pLoF/P); pLoF/P variants restricted to ACMG SF v3.2 genes; and pLoF/P ACMG SF variants retained after expert review according to ACMG/AMP guidelines. Red bars indicate assignable (private) variants; grey indicates the total count. **(E)** Expert-reviewed pathogenic and likely pathogenic variants in ACMG SF genes, plotted at the matrix position of the carrying individual for each of the four 24×24 matrices, and colored by ACMG SF disease category. Gene symbols are shown next to each variant.

## DISCUSSION

In this study, we present a framework for cost-efficient detection of rare genetic variation for population-scale screening. We extended the overlapped pool sequencing approach to 576 individuals in a single batch, automated the labor-intensive DNA pooling steps, improved DNA extraction, and adjusted the library preparation protocol to increase library complexity and facilitate high-coverage sequencing of pools. To improve variant assignment and increase the framework’s robustness to fluctuations in DNA pooling levels, we developed a Bayesian probabilistic model that leverages the redundancy of the two-dimensional pooling design to estimate individual assignment probabilities from initial variant calls and pool-level read evidence. We validated the framework on a reference cohort of individuals with available reference WGS data, achieving a sensitivity comparable to that of individual sequencing, with the additional detection of a clinically relevant *TP53* mosaic variant was missed by the matched 30x WGS. We further applied the framework to a screening cohort of 2,304 apparently healthy anonymous blood donors, yielding variant allele frequency patterns and ACMG SF v.3.2 secondary-finding rates consistent with published estimates.

With the updated sequencing protocol, the framework generated sufficient library complexity and unique-read depth per individual, enabling an increase in the pooling matrix size from 10×10 to 24×24. In the validation cohort, variant detection across target regions had 95% sensitivity compared with individual WGS. For private variants, which are the primary target of the method, sensitivity was 91% (93.5% when excluding QC-flagged low-DNA concentration individuals). This reduction is expected, given the larger pool size and the lower allele fraction of singleton variants, and it remains within the range of cross-method variant concordance and real-world clinical NGS performance^26,27^. In a benchmark of two hereditary cancer panels and WES applied to 24 individuals, pairwise concordance of coding variants between methods ranged from 92.1-93.8%^26^. Finally, because our benchmark used 30x WGS as the reference standard, which itself has imperfect specificity, especially for rare variants and indels^28,29^, the reported numbers likely represent a conservative estimate of the framework’s true performance.

A direct source of reduced sensitivity in our validation experiment was low DNA yield from a subset of dried blood spot cards. Although low-yield extractions would be repeated in a future implementation, we retained these samples in the validation analysis to evaluate the framework’s limits under DNA-input imbalance. This broader range of DNA input also motivated the development of a Bayesian rescue model for private variants with imbalanced read support across their two pools. The model’s largest effects were observed in individuals with low DNA concentration, indicating that it confers additional robustness to the framework. This will also likely be of future importance since DNA extraction and low-concentration measurement are variable^30^, and eliminating the possibility of outliers is difficult at scale.

Even after excluding individuals with low input DNA yield, two pathogenic variants remained unassigned in the validation matrix. This includes a *BRCA2* 5-bp deletion with sufficient coverage at the locus and multiple other variants correctly assigned to the alleged carrier. This indicates that the entire individual was not dropped out, but instead may be due to either a false positive in the matched WGS call set or to differences between the DNA from a recent whole-blood sample used for WGS and the likely more fragmented filter-paper-derived DNA used for DoBSeq. The other variant, a *WT1* 4-bp insertion, was located 5 bp from a 12-bp insertion that was correctly assigned to the same individual. Since pileup data was found for both these insertions, this likely indicates that GATK’s local-reassembly algorithm preferentially resolved one variant over the other.

Although DoBSeq was not initially developed to estimate population frequencies of variants across the entire allele frequency spectrum, the validation experiment provided evidence that within the range possible to model in our data (AF 0–6%), the framework produced allele frequency estimates concordant with those from individual WGS. In the application cohort, the estimated frequencies were similarly consistent with both gnomAD and DanMAC5. Since these cohorts differ in size and diversity, with gnomAD being substantially larger and more heterogeneous than our application cohort, some divergence between estimates is expected. This indicates that DoBSeq could be used as a complement to existing low-cost population-genotyping methods such as SNP chips (DNA microarrays). SNP chips are restricted to a specific set of variants and are widely used to study common genetic variation, but perform poorly when applied to rare variants. In a large-scale evaluation against next-generation sequencing in UK Biobank, SNP-chip genotyping of rare pathogenic *BRCA1* and *BRCA2* variants reached only 34.6% sensitivity and 4.2% positive predictive value^31^. Applying DoBSeq to representative population subsets, in preparation for large-scale screening efforts, could help map allele frequencies for variants in populations or subgroups currently underrepresented in reference databases. Combined with penetrance estimates, these data could help estimate the expected prevalence of corresponding genetic diseases in the population.

The main target of the DoBSeq framework is low-cost identification of candidate carriers of rare pathogenic variants, with confirmation of positive candidates by individual re-sequencing. It is intended for screening purposes in countries with public health or lower healthcare budgets, where individual sequencing of large subsets of the population is not yet economically feasible. Individual sequencing remains more precise and provides more information for each individual. However, when individual sequencing is unavailable, the current alternatives are either no screening or screening via traditional non-genetic methods. HBOC, LS, and FH have been endorsed as reasonable candidates as a starting point for genetic screening of the healthy population, because of their high penetrance and the clinical options available for preventing or mitigating disease in pre-symptomatic individuals^32,33^. For *BRCA1/2*-associated hereditary breast and ovarian cancer, current adult screening practice in most countries relies on family history and select clinical criteria, which are reported to identify only approximately half of *BRCA1/2* pathogenic variant carriers in enriched cohorts and much less in unselected populations^34,35^. Similarly, for FH, screening based on structured clinical criteria, such as the Dutch Lipid Clinic Network criteria, is reported to detect only approximately 30% of confirmed cases^36,37^. At an estimated screening cost of USD 11 per sample, the DoBSeq framework could be a feasible starting point for evaluating genetic screening in large populations.

### Limitations of the study

The DoBSeq framework only detects single-nucleotide variants and small indels. Other classes of genetic variation have been associated with genetic disorders, including structural variants, copy-number variants, and repeat expansions, which would require a range of other methods to cover. The framework is also limited to regions targeted by the capture kit. This could lead to specific regions with lower coverage, such as low-complexity regions, resulting in variants being missed. Furthermore, only variants carried by a single individual in the matrix are assigned, which limits its use for genetic disorders in populations dominated by associated founder variants. This study was also constrained by not having the option to validate any positive samples in the application cohort, limiting the ability to estimate performance in a healthy cohort. The reference cohort was limited to childhood cancer patients, who may carry an unrepresentative distribution of pathogenic variants in cancer predisposition genes, which may lead to a performance that is not representative in other genes and disorders. Consequently, the method requires validation in specific populations for specific genes, variants, and disorders to estimate its true specificity before any clinical application. Finally, this study only addresses the technical aspects of genetic screening. Clinical implementation in a healthy population raises ethical questions about consent, the return of results, and counseling capacity, which will have to be resolved before deployment, regardless of the screening technology.

## METHODS

### Cohorts

For the reference cohort used in model development and validation of the framework, this study included samples from the Danish childhood cancer genomics studies iCOPE and STAGING. The participants were all born in Denmark. WGS was performed on germline DNA as part of the original studies. DoBSeq was performed on a subset of the same participants using DNA from dried blood spots constructed for the purpose of this study. For DoBSeq, we included a total of 776 patients, divided into 3 matrices: two 10×10 matrices, each containing 100 individuals, with data already available from the pilot study described by Stoltze et al.^11^, and a new, larger 24×24 matrix containing 576 individuals. The individuals included in the application part of the study were 2304 anonymized adults from the Danish Blood Donor Study (DBDS) arranged in four 24×24 matrices. No other data or information was available for the participants in this cohort.

### Reference variant calls from individual whole-genome sequencing data

For the reference cohort, matrix T1, V1, and V2, individual germline DNA samples extracted from whole blood were sequenced on an Illumina HiSeqX platform (Illumina, San Diego, CA, USA) using paired-end 150 bp reads with a mean coverage target of 30X. Sequencing reads were mapped to the GRCh38 primary human reference assembly with BWA-MEM2 v2.2.1^38^. Duplicates were marked, and base quality scores recalibrated with GATK v4.5.0.0, following the GATK Best Practices workflow for germline short-variant discovery^39,40^. To limit the analysis of the target regions of the study, alignment data was restricted to the genomic intervals of the custom 582-gene panel using SAMtools v1.20^41^. Joint variant calling for individuals within each matrix was then performed using the standard GATK pipeline: per-sample variant discovery with HaplotypeCaller in GVCF mode, sample aggregation via GenomicsDB import, and joint genotyping with GenotypeGVCFs. Resulting VCFs were normalized using BCFtools v1.20^41^ to left-align indels and split multi-allelic sites.

### Sample handling and overlapped pool sequencing

Frozen whole blood samples from the 576 participants in the STAGING study were thawed, applied to Whatman Specimen 903® Collection Paper, dried for 3 hours, and stored at -20⁰C. Four 3.2 mm punches from the dried blood spots were taken using a Panthera puncher in deep well plates, and DNA was extracted using the Magbio HighPrep Blood & Tissue DNA EP Kit -DX on a Biomek i7. DNA concentrations were measured using the Quant-IT HS kit (Thermo Fischer), and samples were normalized and pooled in 24 rows and 24 columns. Pools were concentrated using ultrafiltration plates (Qiagen). Sequencing libraries from matrix pools were prepared using the Twist Library Preparation EF Kit 1, 2.0 (Twist Bioscience) following the manufacturer’s instructions with the following modifications: 10 min fragmentation, 20 min ligation, and 7 cycles PCR amplification. Capture pools were prepared in 8-plex with 750 ng library per sample and hybridized overnight with Twist Hybridization Reagents using a custom panel (582 genes, 16942 probes, Twist Design ID: TE-93129556, Twist Biosciences) and captured and washed following the manufacturer’s instructions. Libraries were sequenced on a NovaSeq6000.

### Analysis of pool sequencing data and initial assignment of variants

DoBSeq data was analyzed using DoBSeqWF^12^ (v. 0.3.0), our previously published pipeline for overlapped pooled sequencing data. Briefly, sequencing reads were mapped to the GRCh38 primary human reference genome using BWA-MEM2^38^ (v. 2.2.1), and duplicates were marked using GATK MarkDuplicates^42^ (v. 4.5.0.0). Variants were called using GATK HaplotypeCaller^42^ (v. 4.5.0.0) with relevant ploidy settings for the specific matrix configuration (10×10: 20, 24×24: 48).

To provide raw allele-level coverage for the downstream probabilistic model, alignment data in BAM format was converted to mpileup using SAMtools^41^ (v. 1.20). For each genomic position with a variant call in any pool, the number of reads supporting the reference and alternative alleles was extracted from the mpileup files across all pools using a custom Python script.

These per-pool, per-position read counts served as input to the probabilistic models described below.

Finally, as an initial step in the assignment process, variants called in exactly one row pool and one column pool were directly assigned to the individual at the corresponding matrix intersection, as described previously^12^.

### Gene panel design

The custom 582-gene panel was designed to capture genes for which pathogenic variants are associated with medically actionable disorders or conditions considered relevant for genomic screening in the Danish population (see **Table S1**). Gene selection was performed by the multidisciplinary study investigators based on clinical actionability and evidence from established screening initiatives. The panel includes all genes from the ACMG Secondary Findings v3.2 list, the 113-gene hereditary cancer panel used in T1 and V1, genes associated with high-risk childhood cancer predisposition, inherited cardiovascular disorders, and inborn errors of metabolism relevant to current or emerging newborn screening programs. Candidate genes were further informed by comparison with multiple published and ongoing genomic screening initiatives, including the Guardian Study^43^, the UK Generation Study^10^, and panels proposed by Kingsmore^44^, Milko^45^, and Chen^46^, prioritizing genes supported across multiple frameworks for population genomic screening.

### Variant annotation and reference data filtering

Variants from both pool and individual sequencing data were annotated using Ensembl Variant Effect Predictor^47^ (VEP, v. 111.0). This annotation included ClinVar^48^ (release 2023-09-03) clinical significance assertions, gnomAD^29^ (v. 4.1) and danMAC5^17^ (release 2022-10-26) population allele frequencies, and loss-of-function confidence annotations from the LOFTEE plugin^29^.

For the individual WGS reference data, variants were filtered to retain only those with an alternative allele fraction above 0.3, an alternative allele depth of at least 10, and a quality-by-depth (QD) score of at least 2.0. SNVs additionally required allele-specific strand odds ratio (AS_SOR) below 3.0 and allele-specific Fisher strand bias (AS_FS) below 60.0; indels required AS_FS below 200.0.

### Probabilistic model for quantifying variant assignment confidence

To quantify confidence in variant assignment, we developed a Bayesian model to evaluate whether a candidate individual is the only carrier of a given variant within a pooling matrix. For each pool at a variant position, we compared two models using a Bayes factor (BF): a carrier model (M1), in which the pool contains a true variant carrier, and a noise model (M0), in which variant-supporting reads arise from background noise.

For each model, we computed the probability of observing k variant-supporting reads from n total reads using a beta–binomial model:

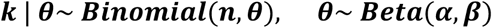

Here, θ represents the underlying probability that a read supports the variant and is treated as a random variable following a Beta distribution with model-specific parameters α and β. The beta-binomial distribution has the following probability mass function:

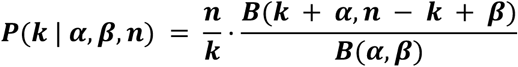

where B denotes the Beta function. Model specific parameters α and β were estimated empirically. For the carrier model (M1), parameters were estimated empirically for each individual and pooling dimension by aggregating variant-supporting reads and coverage from previously assigned variants (and using the theoretical median as a starting point and a prior as regularization “prior_strength”). For the noise model (M0), they were estimated using variant-supporting reads and coverage from all non-candidate pools. The mean of the Beta distribution, α/(α+β), represents the expected variant allele fraction, and α+β controls prior strength. The log Bayes factor comparing the two models was computed as:

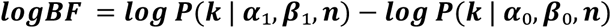

Posterior odds were obtained by combining the log Bayes factor with prior odds:

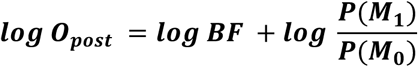

Posterior probabilities were computed using the logistic (sigmoid) function 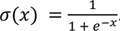. For the candidate pool, the posterior of being a true carrier was:

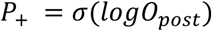

And for each non-candidate pool, the probability of being a non-carrier was:

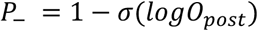

Assuming a single carrier per matrix, the joint probability that a candidate individual is the unique carrier of a variant was computed as:

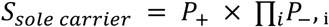

### Allele frequency estimation

In order to estimate the allele frequencies of variants within a matrix, we developed a simple probabilistic model based on the number of positive pools. The model assumes that carriers are heterozygous and uniformly distributed across the *R x C* sample matrix. By design, each carrier is present in one row and one column.

The probability of a single carrier being present in a specific row is 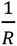, so the probability of not observing that carrier in a given row is 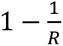. For *h* independent carriers, the probability that none is present in a given row is then 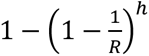. The expected number of positive row pools given *h* carriers in a matrix with R rows is then:

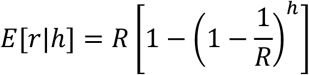

Inverting this expectation gives the carrier count estimated from the number of positive row pools *r*, and similarly by the number of positive column pools *c*:

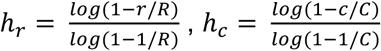

The combined carrier count estimate is the mean of the two:

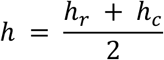

Under the assumption of heterozygous carriers, the allele frequency in the matrix is then:

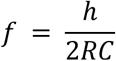

### Comparison of variant calls between reference and pooled data

We performed two comparisons between the pooled sequencing data and the individual WGS reference data, a pool-level detection comparison and a private-variant assignment comparison. For the pool-level detection comparison, the reference variant calls of all individuals assigned to a given pool were merged into a single per-pool VCF using bcftools merge, with duplicate entries removed. This was repeated across all pools in each matrix (20 pools for 10×10 matrices, 48 pools for 24×24 matrices). For each pool, the variants called in the pooled sequencing data were compared to the merged reference set using RTG-tools vcfeval (Cleary et al, 2015, unpublished software, v. 3.13) with the “--squash-ploidy” option.

For the private-variant assignment comparison, the reference variant call set for each individual was intersected against all other individuals in the matrix using bcftools isec to identify variants unique to that individual (100 individuals for 10×10 matrices, 576 individuals for 24×24 matrices). These per-individual private variant sets were then compared to the variants assigned by the pinpointing algorithm similarly using RTG-tools vcfeval.

### Clinical variant review

Variants were classified by a panel of multidisciplinary experts, comprising senior clinical oncogeneticists, molecular biologists, bioinformaticians, and pediatric oncologists. When necessary, experts from other specialties were consulted. Variants were classified as benign (class 1), likely benign (class 2), variant of unknown significance (class 3), likely pathogenic (class 4), or pathogenic (class 5), according to the American College of Medical Genetics and Genomics and the Association for Molecular Pathology guidelines^18^, as recently described^16,49^.

### Statistical analysis

All statistical analyses were performed using the software R (v.4.2.2), with the pROC package for ROC-AUC computation and the Hmisc package for Wilson score 95% confidence intervals for carrier proportions. Correlations between paired allele frequency estimates were estimated using Spearman’s ρ and Pearson’s r, with mean absolute error (MAE) computed as the mean of the absolute differences between paired estimates. The association between pre-pooling DNA concentration and the increase in rescue-model sensitivity was tested using a one-sided Spearman correlation test.

## DECLARATIONS

### Ethics approval and consent to participate

This research was approved by the Capital Region Committee on Health Research Ethics (H-15016782) and the Danish Data Protection Agency (RH-2016-219) and for DBDS it was approved by the Zealand Region Regional Committee on Health Research Ethics (SJ-740) and the Danish Data Protection Agency (RH-99-2019). Samples from DBDS were anonymized.

Written and oral consent was obtained from parents or legal guardians for each participant’s involvement in the study. In accordance with Danish Law, adolescent participants 15 years or older were actively included in the consent process. This meant that while parents/legal guardians legally make the final decision regarding the study participation of any child under 18 years of age, adolescent participants were invited into the informed consent process, ensuring that older children’s perspectives and preferences were heard and that their will was fully considered in the decision. This study adhered to the principles of the Helsinki Declaration.

Participant #O4 in the 24×24 validation matrix provided a signed case-report level consent explicitly permitting inclusion of the specific clinical details relevant to his case.

## DATA AVAILABILITY

The DoBSeq workflow is publicly available at https://github.com/RasmussenLab/DoBSeqWF

(DOI: 10.5281/zenodo.20953988) and the probabilistic model is available at https://github.com/madscort/marbl (DOI: 10.5281/zenodo.21097669). The underlying sequence data cannot be shared publicly due to the privacy of individuals who participated in the study and restrictions from the ethical approval. The data will be shared on reasonable request to the corresponding author, subject to appropriate ethical and legal approvals.

## AUTHOR CONTRIBUTIONS

Conceptualization (STAGING): KS, KW, UKS; Methodology (DoBSeq): JBG, UKS, HH, CMH, SR, KS, KW, TvOH; Methodology (Rescue model): MC, SR; Software: MC; Investigation: MC; Data curation: CMH, UKS, MC; Resources: KW, JBG, MN, CMH, CMJM, AB, MABH, LS, ES, OBVP, CE, SRO; Supervision: SR, UKS, KW; Writing – original draft: MC, CMJM, UKS; Writing – review & editing: All authors.

## ACKNOWLEDGMENTS

Anja Hirche, Solvej Margarete Aldinger Kullegaard, and colleagues for coordination and executions of patient inclusion, sample collection, and data curation for the STAGING/iCOPE study.

