## Supplemental figures for "Low-cost rare variant detection for population scale genetic screening"

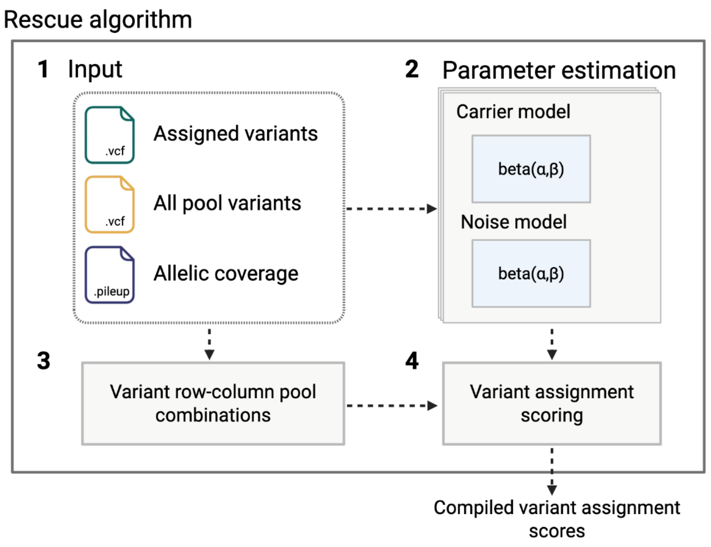


**Figure S1.** Rescue algorithm overview.
(**1**) The algorithm takes initial assigned variants, all variant calls in all pools and the total and alternative allele coverage for all genomic positions. (**2**) Based on the assigned variants and their total and alternative allele coverage, the algorithm estimates alpha and beta parameters of beta distributions specific to each individual in the matrix. (**3**) The algorithm identifies variants called in only one pool and collects pool level alignment information at those positions across the matrix. (**4**) For each single-call variant, candidate assignments are scored under the competing carrier and background beta-binomial models using the individual specific parameters, resulting in a final assignment probability for each variant-candidate pair.

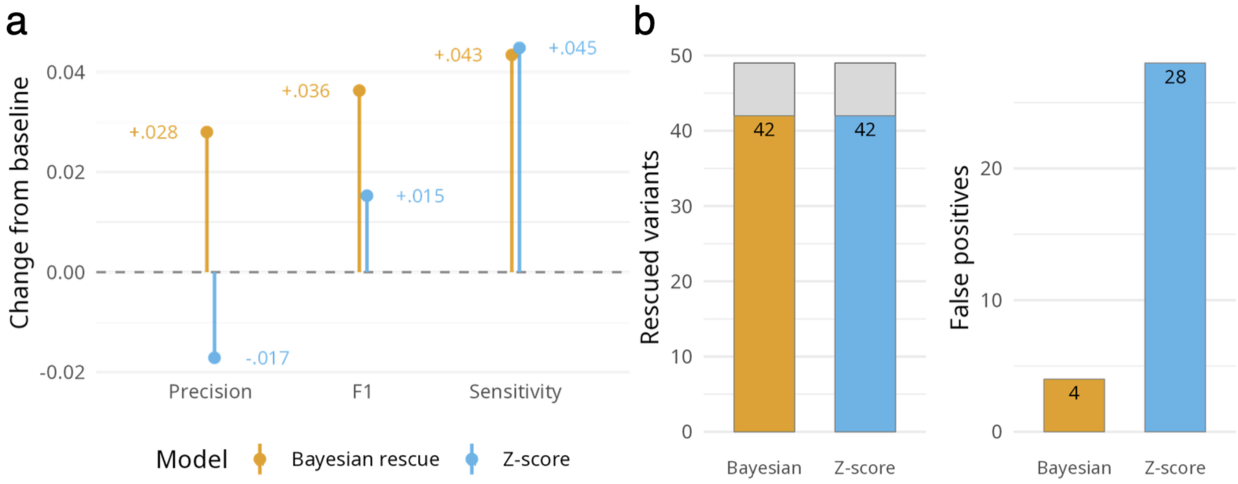


**Figure S2.** Bayesian model validation results on a held-out 10x10 validation matrix (V1). (**A**) Model effect on the performance in assigning private variants to individuals in the 10x10 validation matrix. Changes in performance are shown as rate changes relative to the primary workflow output without a rescue step. (**B**) Number of recovered variants out of the total recoverable variants and the total number of false positive variant assignments for the Bayesian variant rescue model (orange) and the Z-score baseline model (blue). Grey bars indicate the remainder of the recoverable set.

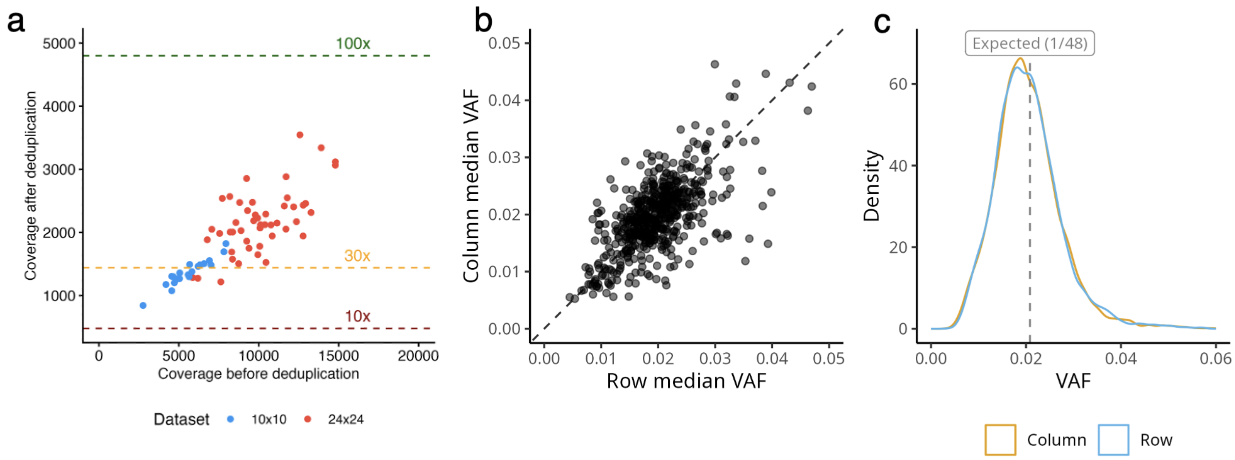


**Figure S3**. Quality control of pooled sequencing data.
(**A**) Coverage per pool before and after deduplication for the 10x10 (V1) and the 24x24 (V2) from the reference cohort. Dashed lines indicate per-allele coverage of 10x, 30x, and 100x, assuming 48 alleles per pool. (**B**) Per-individual median variant allele fraction (VAF) computed across assigned heterozygous private variants, separately for the row pool and column pool. Each point is one individual. The dashed line is y = x. (**C**) Density of VAFs across all assigned heterozygous private variants for row pools (blue) and column pools (orange). The dashed line indicates the expected VAF of 1/48 for a true heterozygous variant in a pool of 24 individuals.

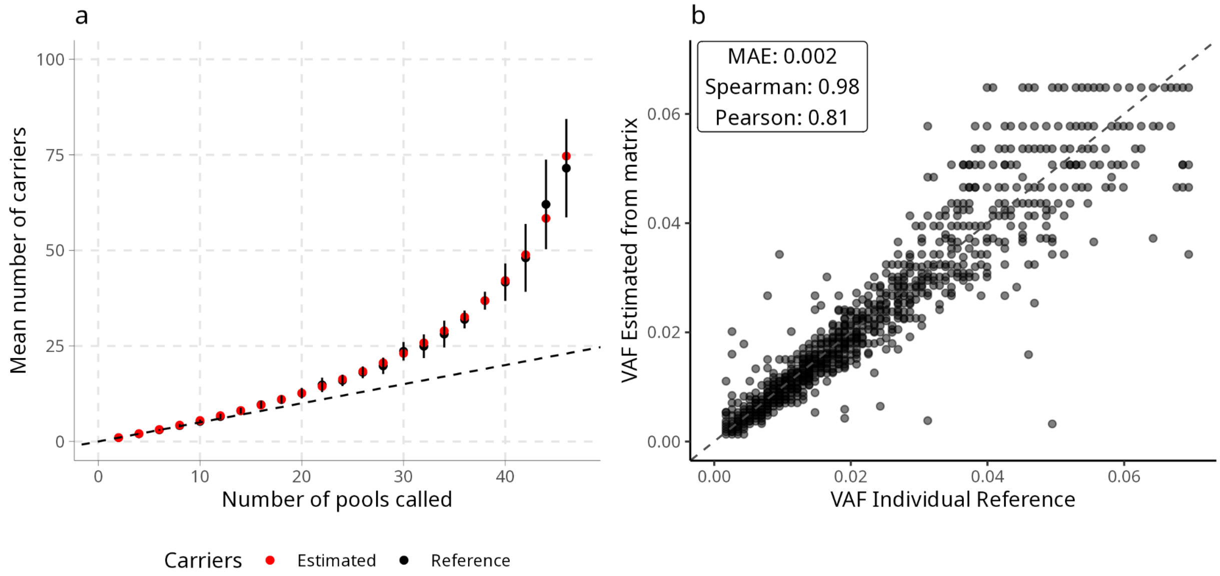


**Figure S4**. Validation of allele frequency estimation model based on data from the 24x24 validation matrix (V2).

(**A**) Mean number of carriers versus number of pools in which the variant was called. Model estimates from pooled data (red) are compared against mean reference counts from individual WGS data (black) shown with standard deviation. The dashed line indicates the theoretical lower bound with one carrier per pool. (**B**) Model estimated VAF from pooled data versus VAF from reference counts.

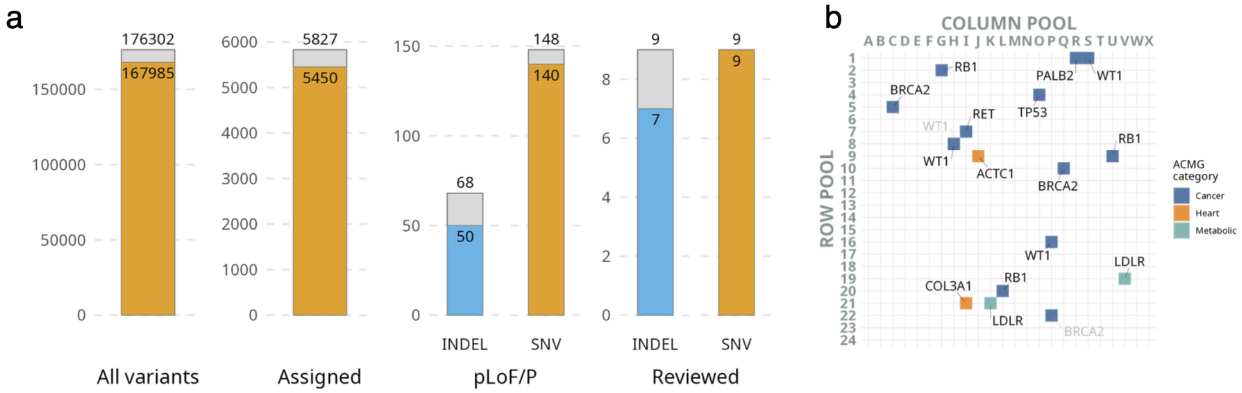


**Figure S5**. Validation of the DoBSeq framework on a batch size of 576 individuals with removal of low DNA yield individuals.

(**A**) Bar charts comparing DoBSeq output (orange) against the individual workflow (grey) at successive filtering stages: all distinct variants (private and common) across the dataset, assigned variants versus private variants, the subset annotated as predicted loss-of-function or ClinVar pathogenic/likely pathogenic (pLoF/P), and expert-reviewed variants within ACMG genes. (**B**) Reviewed variants positioned at their corresponding individual's location in the 24x24 matrix and colored by ACMG category. Variants shown in light grey were identified by individual WGS but not detected by DoBSeq.

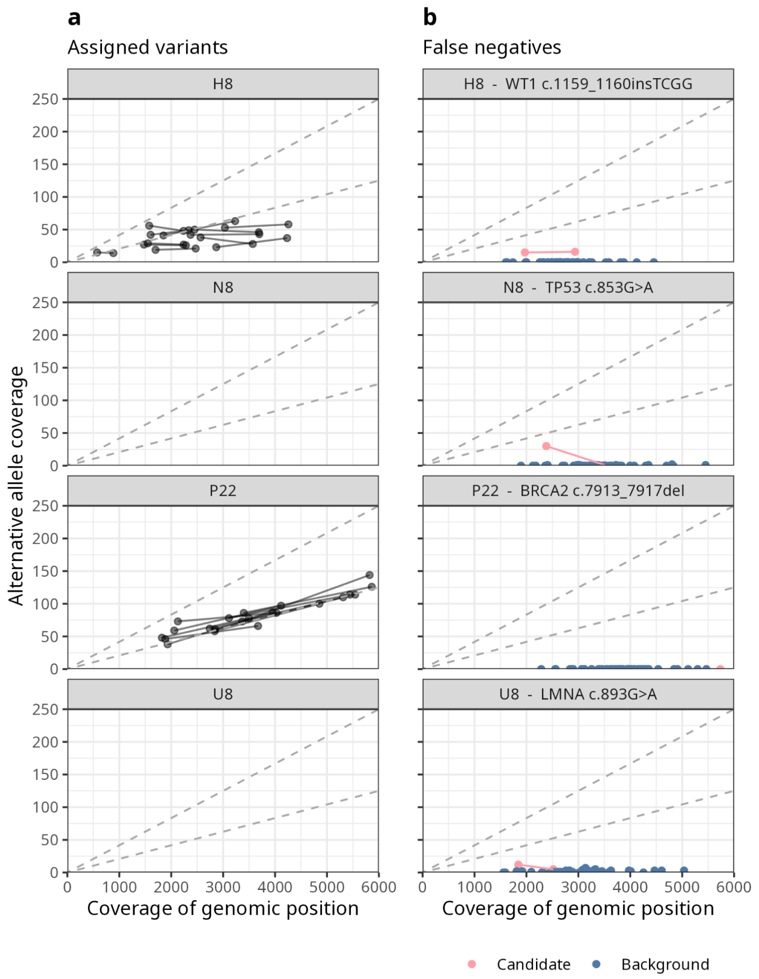


**Figure S6**. Locus coverage plots for clinically relevant variants not assigned by DoBSeq. (**A**) Coverage and alternative allele coverage of all variants assigned to each individual carrying a clinically relevant ACMG SF variant identified by individual WGS, but not assigned by DoBSeq in the 24x24 validation matrix. Each pair of connected dots represents one variant observed in the two pools (row and column). Dashed lines indicate expected alt-allele coverage for homozygous and heterozygous variants, respectively. Individuals #N8 and #U8 had low pre-pooling DNA concentrations and no assigned variants. (**B**) Coverage and alternative allele coverage at the position of the missed variants for the row–column corresponding to each individual (rose) compared with the same position in the remaining 46 pools (blue).

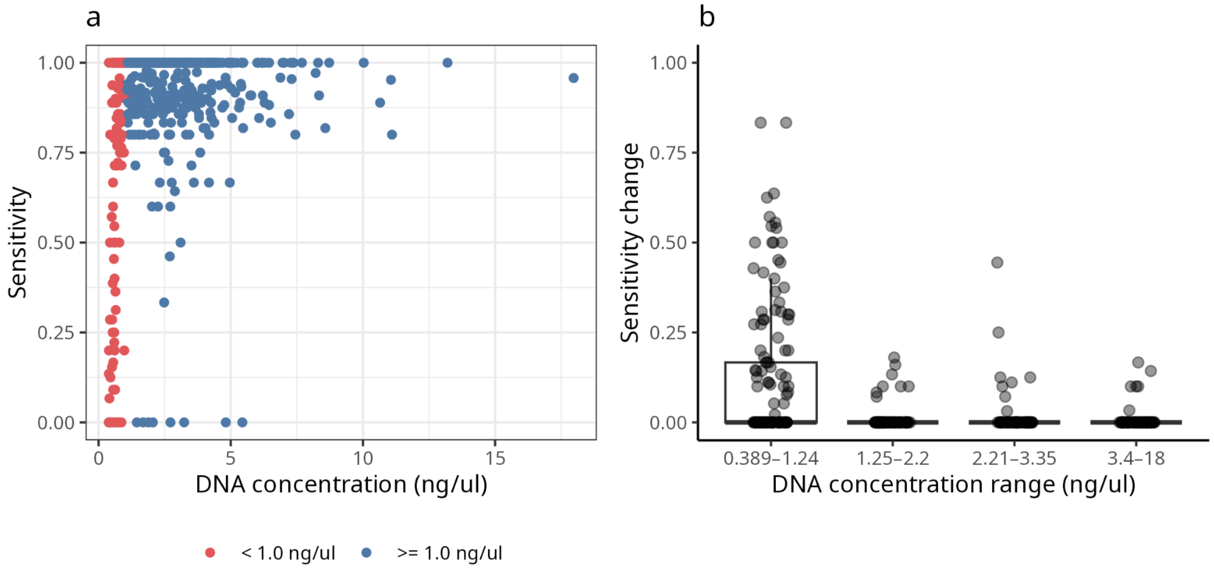


**Figure S7**. Relationship between framework sensitivity and pre-pooling DNA concentrations per individual. (**A**) Per-individual sensitivity of framework for detecting and assigning variants identified by individual WGS, plotted against DNA concentration achieved after extraction from dried bloodspot cards. Equimolar pooling was not achievable for individuals with DNA concentrations below 1 ng/uL (red points). (**B**) Sensitivity gain achieved by the rescue model stratified by pre-pooling DNA concentration (four equal sized bins with ranges shown on x-axis).

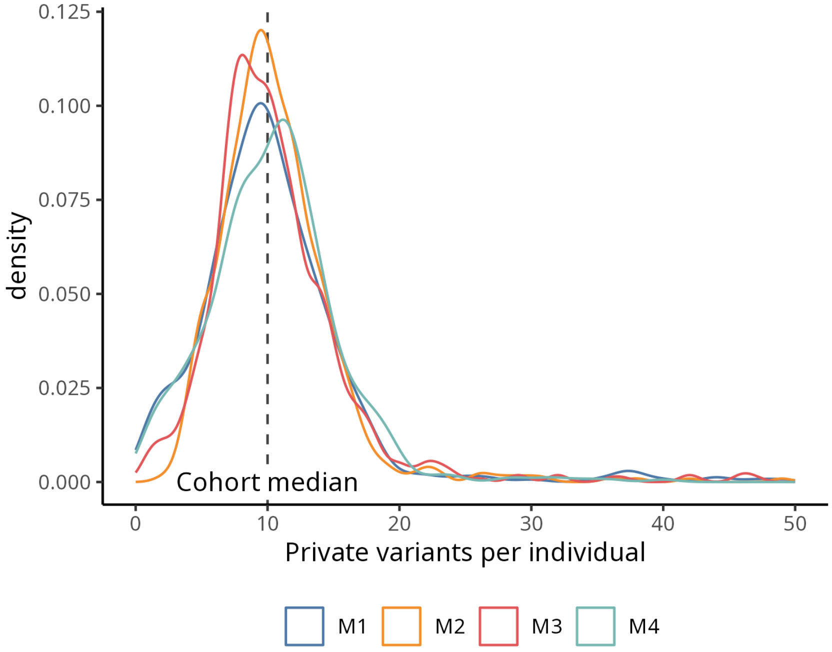


**Figure S8**. Density plot of the number of assigned variants per individual across the four different matrices in the screening cohort. The x-axis is truncated at 50 assigned variants to better represent the data. The truncation removed 15 of 2304 data points (0.65%).

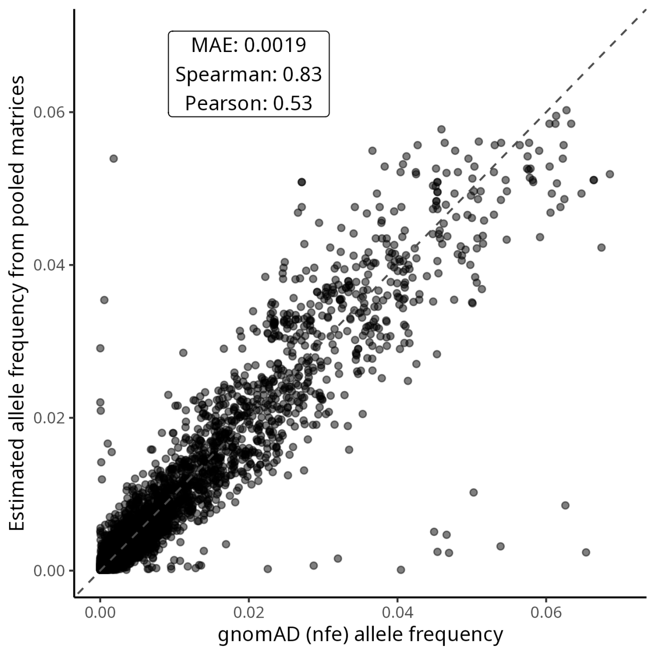


**Figure S9.** Concordance between cohort-level allele frequencies estimated from the four screening matrices (M1-M4) and reference allele frequencies from gnomAD v. 4.1 non-Finnish European subset. Restricted to multi-carrier SNV coding variants present in both datasets.
